# Sex-based differences in the prevalence and determinants of anaemia among children living with HIV Southern Province of Zambia

**DOI:** 10.1101/2025.11.06.25339708

**Authors:** Martin Chakulya, David Chisompola, Lukundo Siame, Benson M. Hamooya, Sepiso K. Masenga

## Abstract

**Background:** anaemia remains a major comorbidity among children living with HIV (CLHIV) in sub-Saharan Africa, yet sex-specific risk factors are poorly characterized. This study investigated the prevalence and sex-based determinants of anaemia among CLHIV in the Southern Province, Zambia.

**Methods:** A retrospective cohort study was conducted using medical records from 321 CLHIV aged 0-14 years. Data on demographic, clinical, and anthropometric variables were analysed. Sex-stratified multivariable logistic regression identified factors associated with anaemia.

**Results:** Overall anaemia prevalence was 47.0% (151/321), with a higher, though not statistically significant, burden in males (52.6%) than females (41.9%). Younger age was a strong, independent risk factor across both sexes. Distinct sex-specific determinants were identified. In males, cotrimoxazole (CTX) use during treatment was associated with increased odds of anaemia (Adjusted Odds Ratio, AOR=3.04; 95% CI: 0.95–9.74). Conversely, among females, the type of caregiver was a significant factor; care provided by an aunt was associated with 90% lower odds of anaemia compared to other arrangements (AOR=0.10; 95% CI: 0.01–0.90). Poor anthropometric indices (height and weight) were significantly associated with anaemia in both sexes.

**Conclusions:** The study findings reveal a high prevalence of anaemia among CLHIV in Zambia, with nuanced sex-based differences in its determinants. The findings advocate for differentiated, gender-sensitive intervention strategies. For boys, careful review of CTX prophylaxis is warranted, while for girls, enhancing supportive caregiving environments may be protective. Integrating these sex-specific approaches into paediatric HIV programs is crucial for reducing the anaemia burden and improving clinical outcomes.

## Introduction

anaemia remains one of the most pervasive and detrimental hematologic complications in children living with Human Immunodeficiency Virus (HIV), persisting as a major contributor to morbidity, impaired neurodevelopment, and diminished quality of life despite global antiretroviral therapy (ART) scale-up [1]. The etology of anaemia in this population is profoundly multifactorial, rooted in the complex interplay of chronic immune activation, nutritional deficiencies, frequent opportunistic infections, and the myelosuppressive effects of ART itself [2,3]. While treatment initiation promotes significant haematological recovery, a substantial burden of anaemia endures, particularly in regions like sub-Saharan Africa where HIV intersects with high rates of malnutrition and other infectious diseases[4,5].

Emerging evidence points to biological sex as a critical modifier of hematologic outcomes, yet this variable remains largely unintegrated into clinical paradigms of paediatric HIV [6,7]. Fundamental sex-based differences in immune function, hormonal regulation, and erythropoietic dynamics are well-documented [8]. The immunomodulatory effects of sex hormones and X-linked genetic factors can influence inflammatory responses, iron metabolism, and erythropoietin sensitivity, creating a divergent biological landscape for the development and progression of anaemia [9,10]. Despite this, paediatric HIV research and care have historically treated children as a homogenous group, potentially obscuring critical sex-specific pathophysiologic pathways and vulnerabilities [11].

This gap is particularly salient in the context of ART. Treatment regimens may exert differential hematologic effects by sex due to variations in drug pharmacokinetics, adherence patterns, and the interaction between ART toxicity and sex-specific physiology [12,13]. The myelosuppression associated with zidovudine, for instance, may manifest differently in boys and girls [2,14]. Furthermore, sociodemographic and nutritional determinants of anaemia such as dietary iron intake and parasitic co-infections are themselves influenced by gendered sociocultural factors, creating a complex web of biological and contextual risk that remains unmapped [15,16].

Elucidating these sex-based disparities is therefore not merely an academic exercise but a necessary step toward precision medicine in paediatric HIV care. A nuanced understanding of how anaemia determinants diverge between boys and girls following ART initiation can refine screening protocols, guide timely nutritional and pharmaceutical interventions, and inform ART regimen selection to optimize haematological recovery. This study aims to address this critical knowledge gap by investigating the sex-specific differences in the prevalence and determinants of anaemia among children living with HIV after ART initiation. By dissecting the intricate interplay of biology, therapy, and context, this research seeks to generate the evidence required for targeted, equitable, and more effective clinical management for all children affected by HIV.

## Methodology

### Design and population

This was a retrospective study conducted among children living with HIV (CLHIV) across 42 health facilities located in 12 districts of the Southern Province, Zambia. Clinical data for participants were abstracted from both paper-based and electronic medical records (SmartCare) at each health facility and entered into the Research Electronic Data Capture (Redcap) system. Information collected included demographic, clinical, and laboratory data such as ART regimen, WHO clinical stage, BMI, duration on ART, retention status, CD4 count, viral load, and pharmacy-related details. In facilities where SmartCare was not fully functional, supplementary data were retrieved from laboratory and pharmacy registers. To ensure accuracy and completeness, data were triangulated between SmartCare and paper-based sources whenever possible. SmartCare is a Zambia Ministry of Health designated, comprehensive electronic health record (EHR) system that facilitates the collection, storage, and reporting of aggregate and patient-level data at facility, district, provincial, and national levels. It provides uniquely identifiable patient information that supports accurate monitoring, evaluation, and informed decision-making. A review of medical records was conducted between November 10 and December 19, 2024.

### Eligibility

We abstracted medical records of children aged below 1 year to 14 years who had a confirmed diagnosis of HIV infection and were enrolled in the antiretroviral therapy (ART) program. Records with missing data on key outcome variables or identified duplicates defined as repeated entries for the same patient within or across facilities based on the unique SmartCare identification number or national registration number were excluded from the analysis.

### Variables in the study

The outcome variable in this study was anaemia among children living with HIV, defined as a haemoglobin concentration below 10.9 g/dL [17].The independent variables included age, sex, health facility, viral load, CD4 count, WHO clinical stage, ART regimen, height, weight, BMI, PMTCT, EMTCT, TB, CTX, and ALT.

### Data collection and sampling

Trained research assistants conducted a comprehensive review of patient records to assemble the demographic and clinical dataset. Data were systematically extracted from two primary sources. The national electronic health record system (SmartCare) and, for facilities not yet integrated into this digital platform, from standardized paper-based registers. To ensure a representative sample, a robust, multi-stage sampling strategy was implemented. First, all eligible health facilities within the study districts were enumerated and assigned a unique identification code. A simple random sample of these facilities was then drawn using a computer-generated random number sequence in Microsoft Excel, guaranteeing an unbiased selection process. Subsequently, the required number of patient files from each selected facility was determined through proportional allocation, weighted by the facility’s volume of paediatric ART clients. This approach ensured that the sample accurately reflected the distribution of the patient population across the health system. Within each facility, individual client files were chosen for inclusion via systematic random sampling. Enrolled participants were followed longitudinally through their scheduled clinic visits, with data captured from successive entries in their medical records (either SmartCare or paper-based). Follow-up continued for each participant until the administrative conclusion of the study or until the occurrence of a censoring event (loss to follow-up, transfer to a non-study facility, or death). All captured data were securely entered and managed using the Research Electronic Data Capture (Redcap) platform to ensure data integrity and security.

### Sample size estimation

From a total of 1,019 medical records available for abstraction, 321 records met the inclusion criteria and were included in the final analysis. A total of 698 records were excluded due to incomplete or missing information, including key demographic variables such as age, sex, and caregiver details, as well as missing clinical data on diagnosis or anaemia outcomes.

**Fig 1.**
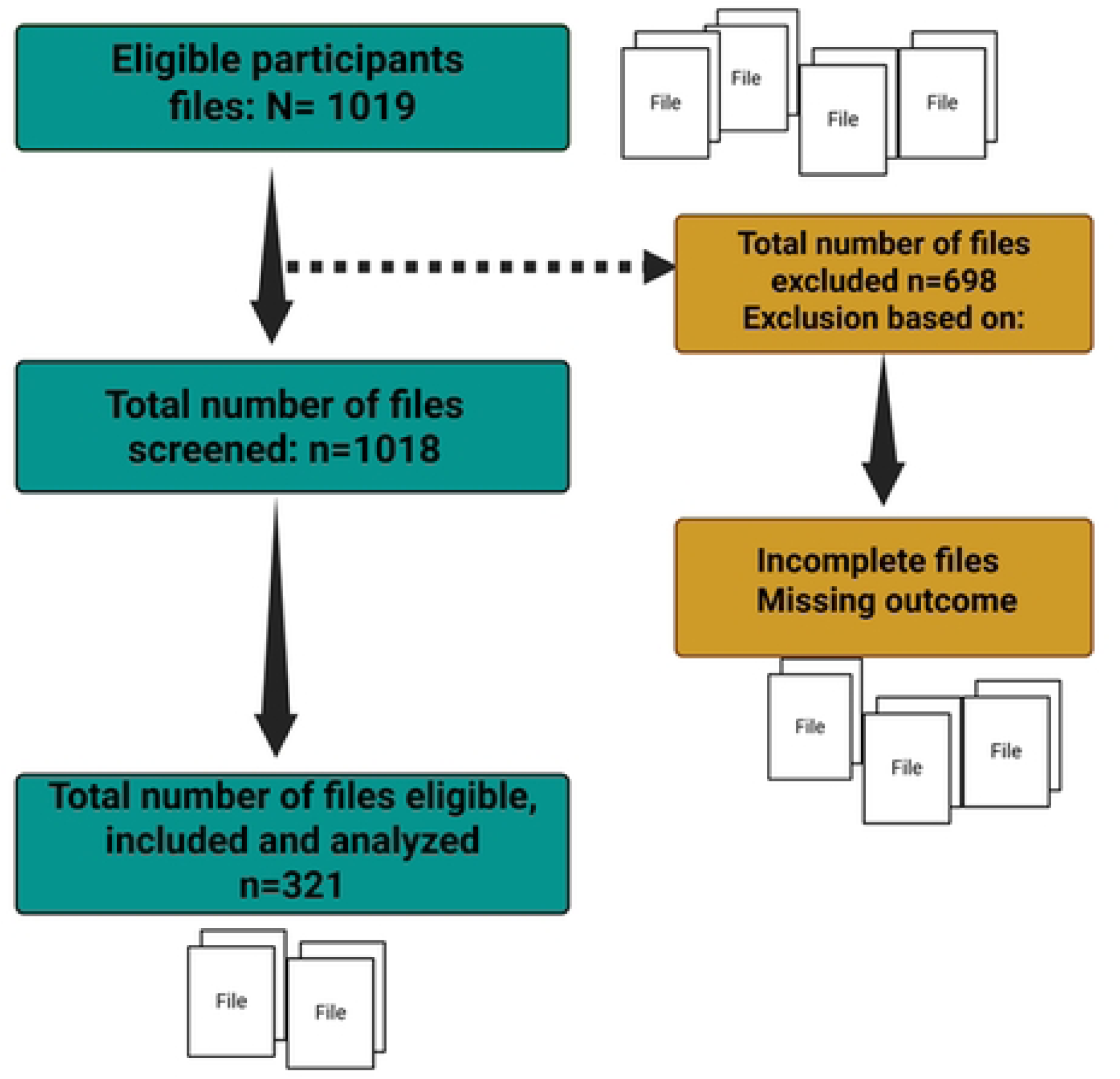
Sample size flow chart.

### Data analysis

We exported data from the REDCap application to Microsoft Excel for cleaning and thereafter analysed in statcrunch. Data were described using frequency and percentages for categorical variables and medians (interquartile ranges) for continuous variables. To test for normality, the Shapiro–Wilk test was used. The Wilcoxon rank-sum test was used to ascertain the statistical difference between the two medians. A relationship between two categorical variables was determined using a chi-squared test. Multivariable logistic regression was used to examine the factors associated with anaemia in PLHIV. The variables age, sex, facility location, WHO stage, ART regimen, and BMI in the adjusted analysis were selected based on previous literature and knowledge from HIV treatment and management experts. Adjusted logistic regression (xtlogit model) was fitted to determine factors associated with anaemia in HIV among paediatrics. The p-value of <0.05 was considered statistically significant.

### Ethical considerations and consent to participants

Ethical approval for the data used in this study was obtained from the Mulungushi University School of Medicine and Health Sciences (SOHMS) Research Ethics Committee (ethics Reference number SMHS-MU2-2025-71) on 14^th^ February 2025.

No information leading to identification of patients during and after analysis was abstracted and entered in the data collection form. Secondary data were used in this project. A written/verbal consent was not applicable and was therefore waived by the ethics committee. We used the Strengthening the Reporting of Observational Studies in Epidemiology (STROBE) checklist to guide reporting, Supplementary file S1.

### Characteristics of the study population

Table 1 presents the relationship between anaemia and various demographic and clinical characteristics among 321 children living with HIV. Overall, 47.0% (n=151) of participants were anaemic, while 53.0% (n=170) were not. The median age of children with anaemia was significantly lower than that of non-anaemic participants (8 years [IQR: 6–11] vs. 11 years [IQR: 8–13]; p < 0.0001), suggesting that younger children were more susceptible to anaemia. Although anaemia was more common among males (52.6%) than females (41.9%), this difference did not reach statistical significance (p = 0.055). ART regimen type demonstrated a significant association with anaemia (p = 0.026). Children on protease inhibitor (PI)-based regimens had the highest anaemia prevalence (57.9%), followed by those on NNRTI/NRTI-based regimens (41.6%), “other” regimens (43.3%), and INSTI-based regimens (35.7%). Similarly, WHO clinical stage was significantly associated with anaemia (p = 0.033); prevalence increased with disease severity, from 40.5% in stage 1 to 67.7% in stage 3, indicating that advanced HIV disease was linked to higher anaemia burden. Geographical location was not significantly associated with anaemia (p = 0.082), although urban children had a higher prevalence (50.7%) than those from rural areas (40.7%). Children under maternal care exhibited a notably higher anaemia rate (52.2%) compared to those with non-maternal caregivers (34.7%). Conversely, care provided by an aunt was associated with a significantly lower prevalence of anaemia (10.5%) than other care arrangements (49.3%). No significant association with anaemia was found for other caregiver types (fathers, grandparents, uncles, siblings, or non-relatives). Furthermore, maternal participation in Prevention of Mother-to-Child Transmission (PMTCT) programs was significantly linked to anaemia in the children. Children whose mothers took antiretroviral medications during pregnancy had a higher anaemia prevalence (54.7%) than those whose mothers did not (50.6%) or whose status was unsure (27.0%). A history of tuberculosis (TB) was inversely associated with anaemia (p = 0.023); only 15.4% of children with a prior TB diagnosis were anaemic compared to 49.5% without such history. Cotrimoxazole (CTX) prophylaxis at ART initiation did not show a significant association (p = 0.131), though anaemia was slightly higher among those on CTX (52.3%) compared to non-recipients (43.8%). Similar patterns were observed for CTX use at any point during treatment (p = 0.094) and before ART initiation (p = 0.437). Anthropometric measures revealed that anaemic children were significantly shorter and lighter than non-anaemic peers. Median height was 110 cm (IQR: 86–156) among anaemic participants and 126 cm (IQR: 105–155) among non-anaemic ones (p = 0.0016), while median weight was 14 kg (IQR: 9–20) versus 20 kg (IQR: 13–25), respectively (p < 0.0001). No significant differences were found in body mass index (p = 0.922), CD4 count (p = 0.601), viral load (p = 0.403), or alanine aminotransferase (ALT) levels (p = 0.611). However, serum creatinine levels were significantly lower among anaemic children (36 µmol/L [IQR: 26–42]) compared to non-anaemic children (41.1 µmol/L [IQR: 30–56]; p = 0.019). Overall, younger age, advanced WHO stage, PI-based ART regimen, maternal caregiving, maternal PMTCT participation, lower anthropometric indices, and serum creatinine levels were significantly associated with anaemia among children living with HIV.

**Table 1.**
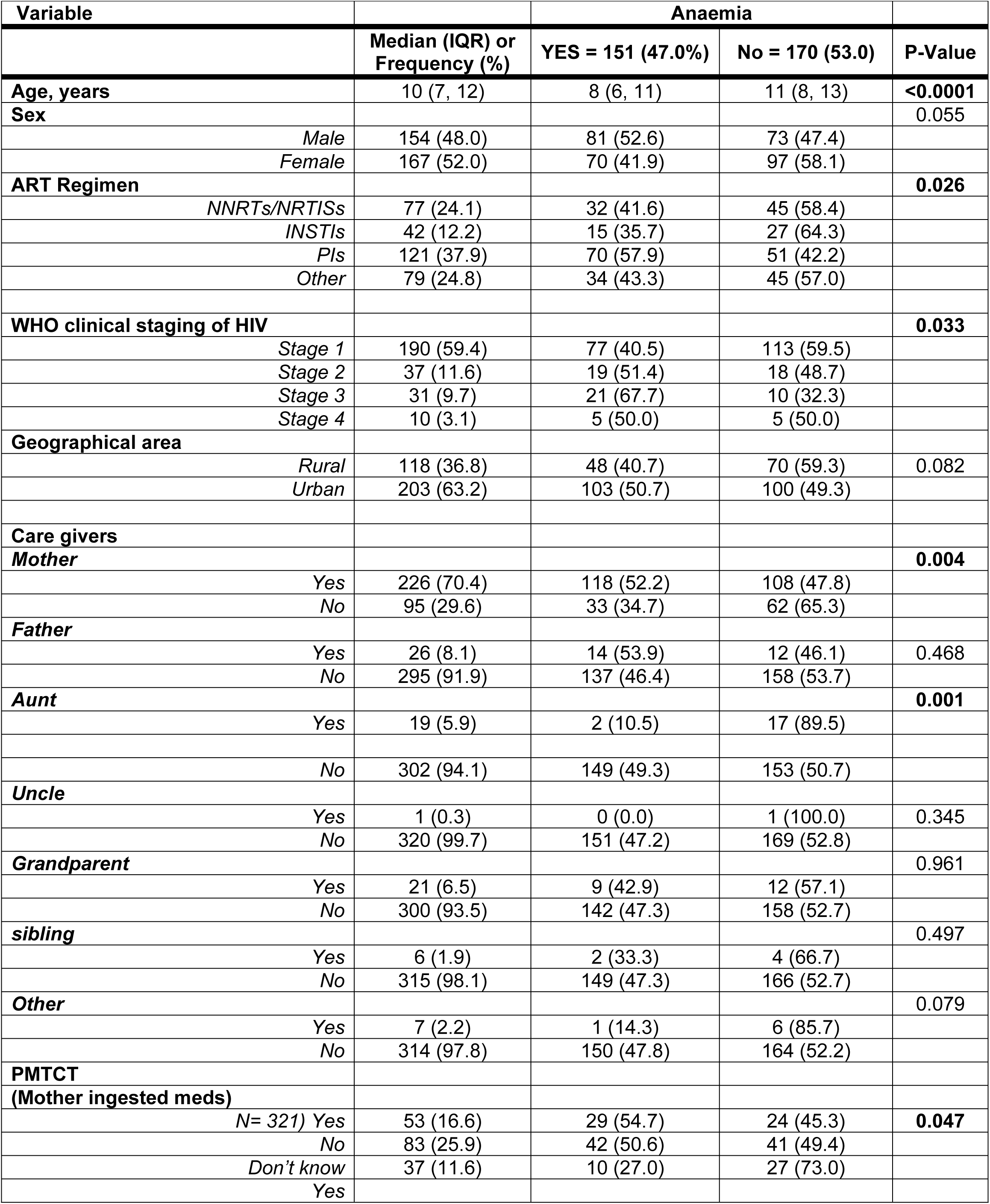
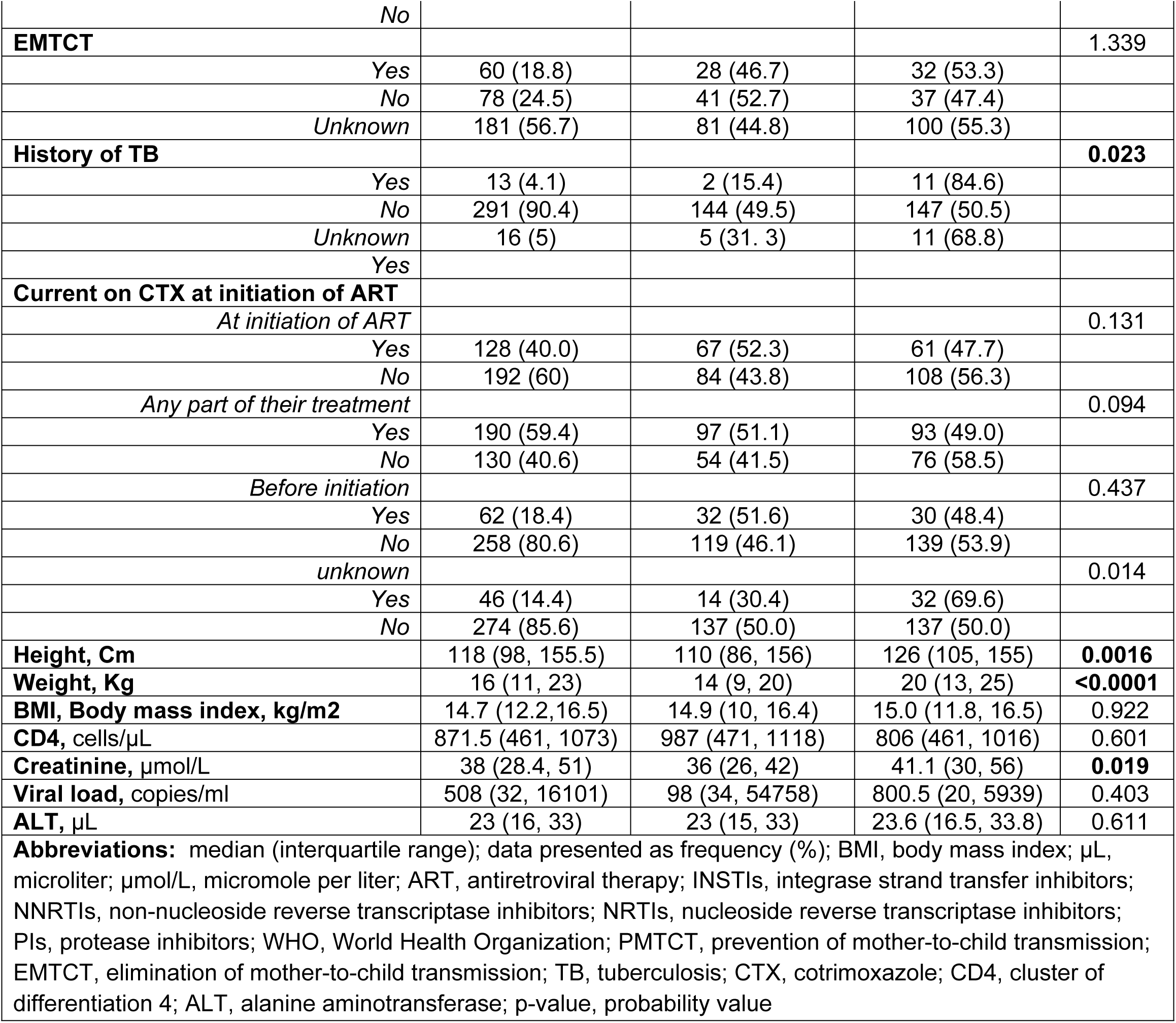
Relationship between demographic and clinical characteristics and Anaemia.

### Comparison between male and female demographic and characteristic of Anaemia in children

The median age of male participants was 9.5 years (IQR: 6–12), while for female participants it was 10 years (IQR: 7–12). Age was significantly associated with anaemia in both sexes (p < 0.0001), with younger children more likely to be anaemic. Regarding ART regimens, the majority of participants were on PIs or NNRTI/NRTI- based therapy. Among males, anaemia prevalence did not differ significantly by ART regimen (p = 0.251). In females, although a higher proportion of anaemic children were on PIs (54.7%) and other regimens (61.8%) compared to NNRTIs (32.6%) or INSTIs (30.0%), this association was not statistically significant (p = 0.061). Most participants were classified as WHO Stage I at baseline, with 58.2% of males and 60.5% of females in this stage. There was no significant association between WHO stage and anaemia in either sex (p = 0.089 in males; p = 0.182 in females). Geographically, most participants resided in rural areas (64.3% of males and 62.3% of females), and residency was not significantly associated with anaemia in either sex (p = 0.516 in males; p = 0.080 in females). Regarding caregivers, most male participants were cared for by their mothers (73.4%), while among females, 67.7% had maternal support. Among females, children cared for by their mothers had a significantly higher prevalence of anaemia (48.7% vs. 27.8%; p = 0.010), whereas in males, caregiver type was not significantly associated with anaemia. Notably, female children under the care of an aunt had a lower prevalence of anaemia (6.7%) compared to those not cared for by an aunt (45.4%; p = 0.004), while no significant differences were observed in males. Analysis of PMTCT interventions revealed that among females whose mothers ingested PMTCT medication type 1, anaemia prevalence was significantly higher (65.4%) compared to other types (p = 0.035). No significant associations were observed in males. EMTCT exposure and history of TB were not significantly associated with anaemia in either sex. Current use of CTX at ART initiation and as part of treatment was not significantly associated with anaemia in both males and females (p > 0.05). Similarly, pre-initiation CTX status and unknown CTX status were not significantly associated with anaemia. Anthropometric measurements showed that male children with anaemia had significantly lower median height (98 cm vs. 121.5 cm; p = 0.0004) and weight (14 kg vs. 20 kg; p = 0.0001) compared to non-anaemic males. Female participants with anaemia also had significantly lower median height (13 cm vs. 19.5 cm; p = 0.0006) and weight (14 kg vs. 20 kg; p = 0.0002) compared to non-anaemic females. No significant differences were observed in BMI, CD4 count, creatinine, viral load, or ALT based on anaemia status in either sex, except that male anaemic children had slightly lower creatinine (31 µmol/L vs. 41 µmol/L; p = 0.0051). See table 2

**Table 2.**
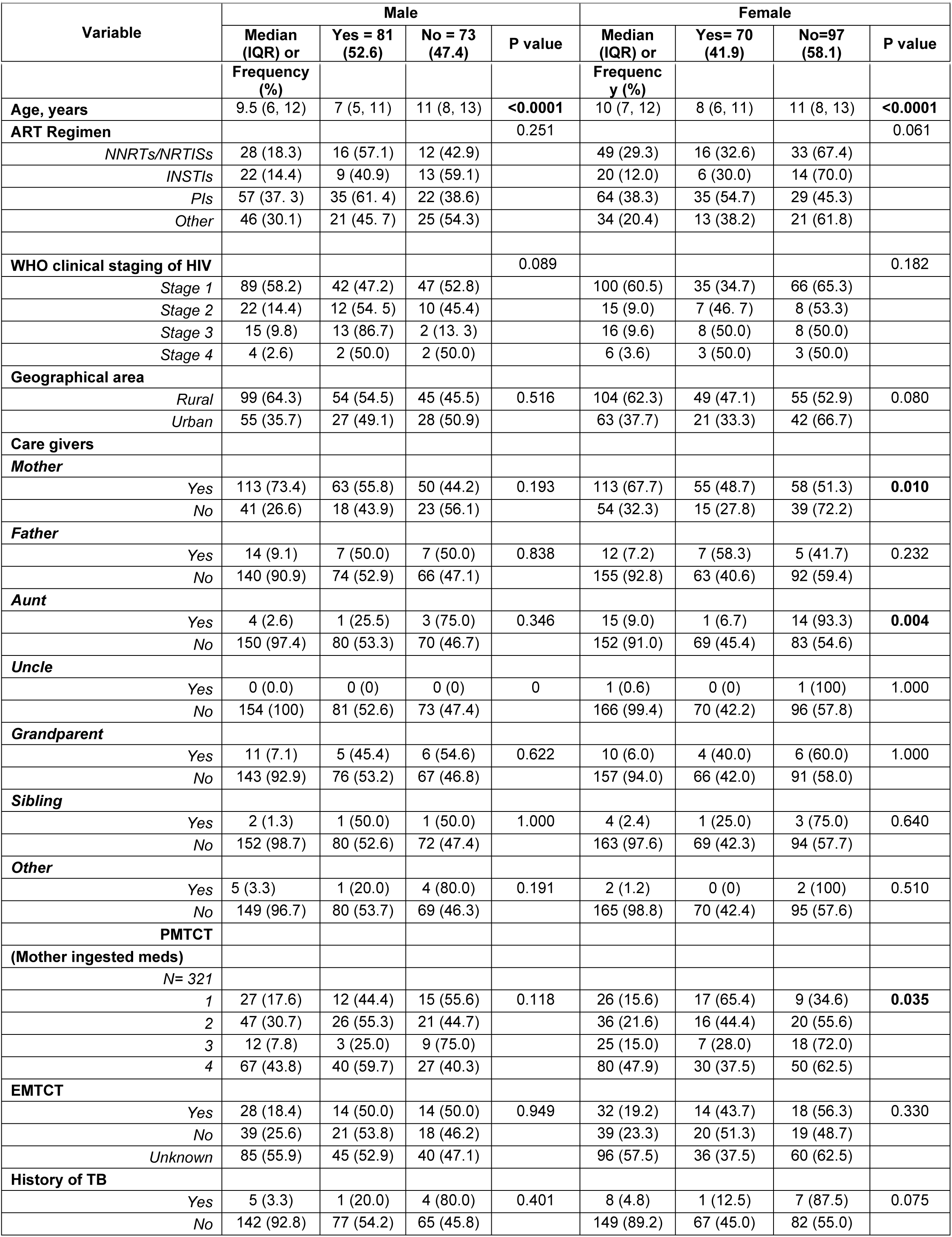
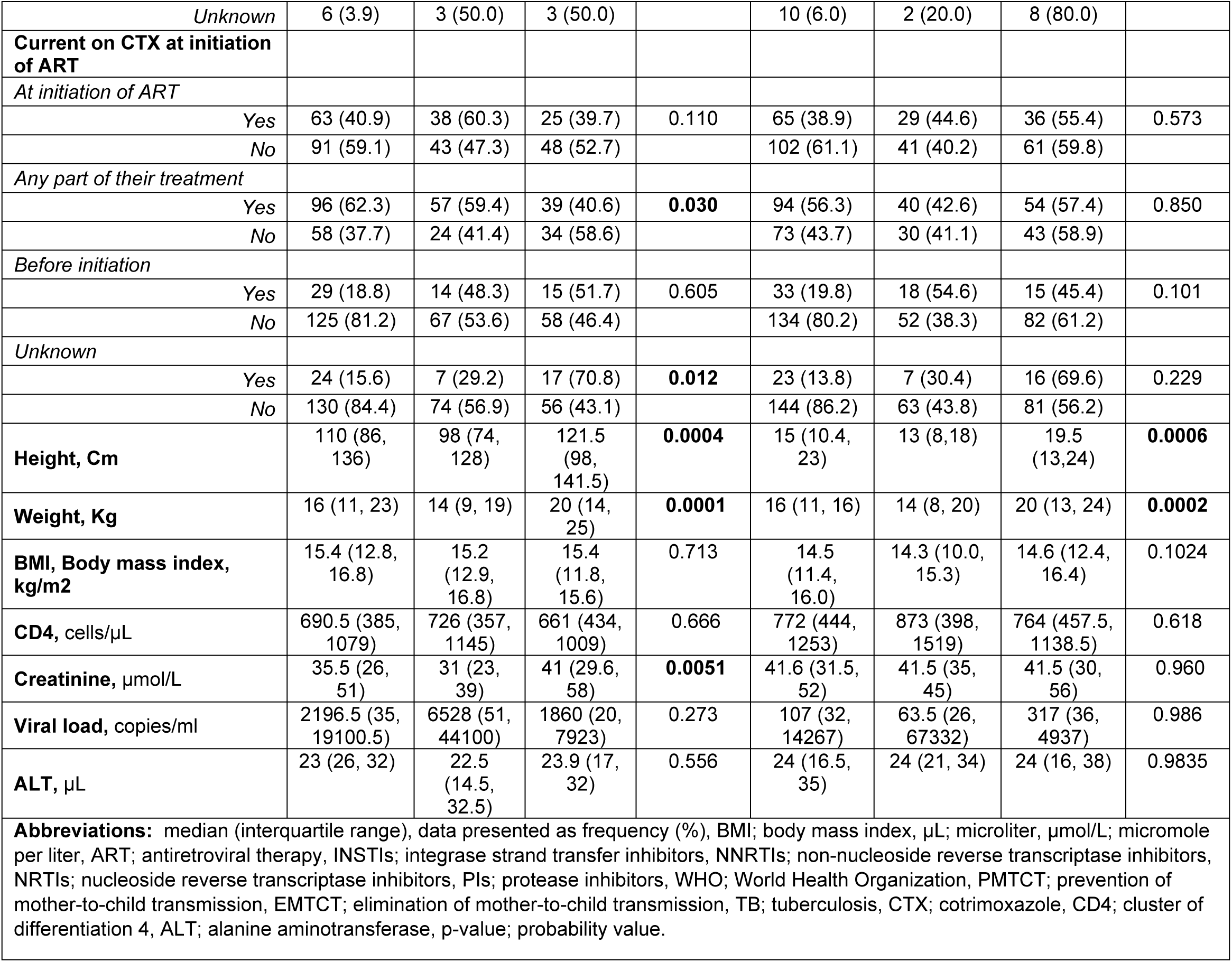
Comparison between male and female demographic and characteristic of Anaemia in children.

|  |  |  |  |  |  |  |  |  |
| --- | --- | --- | --- | --- | --- | --- | --- | --- |
| Variable | Male |  |  |  | Female |  |  |  |
|  | Median (IQR) or | Yes = 81 (52.6) | No = 73 (47.4) | P value | Median (IQR) or | Yes= 70 (41.9) | No=97 (58.1) | P value |

|  | Frequency (%) |  |  |  | Frequency (%) |  |  |  |
| --- | --- | --- | --- | --- | --- | --- | --- | --- |
| **Age, years** | 9.5 (6, 12) | 7 (5, 11) | 11 (8, 13) | **<0.0001** | 10 (7, 12) | 8 (6, 11) | 11 (8, 13) | **<0.0001** |
| **ART Regimen** |  |  |  | 0.251 |  |  |  | 0.061 |
| *NNRTs/NRTIs* | 28 (18.3) | 16 (57.1) | 12 (42.9) |  | 49 (29.3) | 16 (32.6) | 33 (67.4) |  |
| *INSTIs* | 22 (14.4) | 9 (40.9) | 13 (59.1) |  | 20 (12.0) | 6 (30.0) | 14 (70.0) |  |
| *PIs* | 57 (37.3) | 35 (61.4) | 22 (38.6) |  | 64 (38.3) | 35 (54.7) | 29 (45.3) |  |
| *Other* | 46 (30.1) | 21 (45.7) | 25 (54.3) |  | 34 (20.4) | 13 (38.2) | 21 (61.8) |  |
| **WHO clinical staging of HIV** |  |  |  | 0.089 |  |  |  | 0.182 |
| *Stage 1* | 89 (58.2) | 42 (47.2) | 47 (52.8) |  | 100 (60.5) | 35 (34.7) | 66 (65.3) |  |
| *Stage 2* | 22 (14.4) | 12 (54.5) | 10 (45.4) |  | 15 (9.0) | 7 (46.7) | 8 (53.3) |  |
| *Stage 3* | 15 (9.8) | 13 (86.7) | 2 (13.3) |  | 16 (9.6) | 8 (50.0) | 8 (50.0) |  |
| *Stage 4* | 4 (2.6) | 2 (50.0) | 2 (50.0) |  | 6 (3.6) | 3 (50.0) | 3 (50.0) |  |
| **Geographical area** |  |  |  |  |  |  |  |  |
| *Rural* | 99 (64.3) | 54 (54.5) | 45 (45.5) | 0.516 | 104 (62.3) | 49 (47.1) | 55 (52.9) | 0.080 |
| *Urban* | 55 (35.7) | 27 (49.1) | 28 (50.9) |  | 63 (37.7) | 21 (33.3) | 42 (66.7) |  |
| **Care givers** |  |  |  |  |  |  |  |  |
| **Mother** |  |  |  |  |  |  |  |  |
| *Yes* | 113 (73.4) | 63 (55.8) | 50 (44.2) | 0.193 | 113 (67.7) | 55 (48.7) | 58 (51.3) | **0.010** |
| *No* | 41 (26.6) | 18 (43.9) | 23 (56.1) |  | 54 (32.3) | 15 (27.8) | 39 (72.2) |  |
| **Father** |  |  |  |  |  |  |  |  |
| *Yes* | 14 (9.1) | 7 (50.0) | 7 (50.0) | 0.838 | 12 (7.2) | 7 (58.3) | 5 (41.7) | 0.232 |
| *No* | 140 (90.9) | 74 (52.9) | 66 (47.1) |  | 155 (92.8) | 63 (40.6) | 92 (59.4) |  |
| **Aunt** |  |  |  |  |  |  |  |  |
| *Yes* | 4 (2.6) | 1 (25.5) | 3 (75.0) | 0.346 | 15 (9.0) | 1 (6.7) | 14 (93.3) | **0.004** |
| *No* | 150 (97.4) | 80 (53.3) | 70 (46.7) |  | 152 (91.0) | 69 (45.4) | 83 (54.6) |  |
| **Uncle** |  |  |  |  |  |  |  |  |
| *Yes* | 0 (0.0) | 0 (0) | 0 (0) | 0 | 1 (0.6) | 0 (0) | 1 (100) | 1.000 |
| *No* | 154 (100) | 81 (52.6) | 73 (47.4) |  | 166 (99.4) | 70 (42.2) | 96 (57.8) |  |
| **Grandparent** |  |  |  |  |  |  |  |  |
| *Yes* | 11 (7.1) | 5 (45.4) | 6 (54.6) | 0.622 | 10 (6.0) | 4 (40.0) | 6 (60.0) | 1.000 |
| *No* | 143 (92.9) | 76 (53.2) | 67 (46.8) |  | 157 (94.0) | 66 (42.0) | 91 (58.0) |  |
| **Sibling** |  |  |  |  |  |  |  |  |
| *Yes* | 2 (1.3) | 1 (50.0) | 1 (50.0) | 1.000 | 4 (2.4) | 1 (25.0) | 3 (75.0) | 0.640 |
| *No* | 152 (98.7) | 80 (52.6) | 72 (47.4) |  | 163 (97.6) | 69 (42.3) | 94 (57.7) |  |
| **Other** |  |  |  |  |  |  |  |  |
| *Yes* | 5 (3.3) | 1 (20.0) | 4 (80.0) | 0.191 | 2 (1.2) | 0 (0) | 2 (100) | 0.510 |
| *No* | 149 (96.7) | 80 (53.7) | 69 (46.3) |  | 165 (98.8) | 70 (42.4) | 95 (57.6) |  |
| **PMTCT** |  |  |  |  |  |  |  |  |
| **(Mother ingested meds)** |  |  |  |  |  |  |  |  |
| *N= 321* |  |  |  |  |  |  |  |  |
| *1* | 27 (17.6) | 12 (44.4) | 15 (55.6) | 0.118 | 26 (15.6) | 17 (65.4) | 9 (34.6) | **0.035** |
| *2* | 47 (30.7) | 26 (55.3) | 21 (44.7) |  | 36 (21.6) | 16 (44.4) | 20 (55.6) |  |
| *3* | 12 (7.8) | 3 (25.0) | 9 (75.0) |  | 25 (15.0) | 7 (28.0) | 18 (72.0) |  |
| *4* | 67 (43.8) | 40 (59.7) | 27 (40.3) |  | 80 (47.9) | 30 (37.5) | 50 (62.5) |  |
| **EMTCT** |  |  |  |  |  |  |  |  |
| *Yes* | 28 (18.4) | 14 (50.0) | 14 (50.0) | 0.949 | 32 (19.2) | 14 (43.7) | 18 (56.3) | 0.330 |
| *No* | 39 (25.6) | 21 (53.8) | 18 (46.2) |  | 39 (23.3) | 20 (51.3) | 19 (48.7) |  |
| *Unknown* | 85 (55.9) | 45 (52.9) | 40 (47.1) |  | 96 (57.5) | 36 (37.5) | 60 (62.5) |  |
| **History of TB** |  |  |  |  |  |  |  |  |
| *Yes* | 5 (3.3) | 1 (20.0) | 4 (80.0) | 0.401 | 8 (4.8) | 1 (12.5) | 7 (87.5) | 0.075 |
| *No* | 142 (92.8) | 77 (54.2) | 65 (45.8) |  | 149 (89.2) | 67 (45.0) | 82 (55.0) |  |
| *Unknown* | 6 (3.9) | 3 (50.0) | 3 (50.0) |  | 10 (6.0) | 2 (20.0) | 8 (80.0) |  |
| **Current on CTX at initiation of ART** |  |  |  |  |  |  |  |  |
| *At initiation of ART* |  |  |  |  |  |  |  |  |
| Yes | 63 (40.9) | 38 (60.3) | 25 (39.7) | 0.110 | 65 (38.9) | 29 (44.6) | 36 (55.4) | 0.573 |
| No | 91 (59.1) | 43 (47.3) | 48 (52.7) |  | 102 (61.1) | 41 (40.2) | 61 (59.8) |  |
| *Any part of their treatment* |  |  |  |  |  |  |  |  |
| Yes | 96 (62.3) | 57 (59.4) | 39 (40.6) | **0.030** | 94 (56.3) | 40 (42.6) | 54 (57.4) | 0.850 |
| No | 58 (37.7) | 24 (41.4) | 34 (58.6) |  | 73 (43.7) | 30 (41.1) | 43 (58.9) |  |
| *Before initiation* |  |  |  |  |  |  |  |  |
| Yes | 29 (18.8) | 14 (48.3) | 15 (51.7) | 0.605 | 33 (19.8) | 18 (54.6) | 15 (45.4) | 0.101 |
| No | 125 (81.2) | 67 (53.6) | 58 (46.4) |  | 134 (80.2) | 52 (38.3) | 82 (61.2) |  |
| *Unknown* |  |  |  |  |  |  |  |  |
| Yes | 24 (15.6) | 7 (29.2) | 17 (70.8) | **0.012** | 23 (13.8) | 7 (30.4) | 16 (69.6) | 0.229 |
| No | 130 (84.4) | 74 (56.9) | 56 (43.1) |  | 144 (86.2) | 63 (43.8) | 81 (56.2) |  |
| **Height, Cm** | 110 (86, 136) | 98 (74, 128) | 121.5 (98, 141.5) | **0.0004** | 15 (10.4, 23) | 13 (8, 18) | 19.5 (13, 24) | **0.0006** |
| **Weight, Kg** | 16 (11, 23) | 14 (9, 19) | 20 (14, 25) | **0.0001** | 16 (11, 16) | 14 (8, 20) | 20 (13, 24) | **0.0002** |
| **BMI, Body mass index, kg/m<sup>2</sup>** | 15.4 (12.8, 16.8) | 15.2 (12.9, 16.8) | 15.4 (11.8, 15.6) | 0.713 | 14.5 (11.4, 16.0) | 14.3 (10.0, 15.3) | 14.6 (12.4, 16.4) | 0.1024 |
| **CD4, cells/μL** | 690.5 (385, 1079) | 726 (357, 1145) | 661 (434, 1009) | 0.666 | 772 (444, 1253) | 873 (398, 1519) | 764 (457.5, 1138.5) | 0.618 |
| **Creatinine, μmol/L** | 35.5 (26, 51) | 31 (23, 39) | 41 (29.6, 58) | **0.0051** | 41.6 (31.5, 52) | 41.5 (35, 45) | 41.5 (30, 56) | 0.960 |
| **Viral load, copies/ml** | 2196.5 (35, 19100.5) | 6528 (51, 44100) | 1860 (20, 7923) | 0.273 | 107 (32, 14267) | 63.5 (26, 67332) | 317 (36, 4937) | 0.986 |
| **ALT, μL** | 23 (26, 32) | 22.5 (14.5, 32.5) | 23.9 (17, 32) | 0.556 | 24 (16.5, 35) | 24 (21, 34) | 24 (16, 38) | 0.9835 |
| **Abbreviations:** median (interquartile range), data presented as frequency (%), BMI; body mass index, μL; microliter, μmol/L; micromole per liter, ART; antiretroviral therapy, INSTIs; integrase strand transfer inhibitors, NNRTIs; non-nucleoside reverse transcriptase inhibitors, NRTIs; nucleoside reverse transcriptase inhibitors, PIs; protease inhibitors, WHO; World Health Organization, PMTCT; prevention of mother-to-child transmission, EMTCT; elimination of mother-to-child transmission, TB; tuberculosis, CTX; cotrimoxazole, CD4; cluster of differentiation 4, ALT; alanine aminotransferase, p-value; probability value. | | | | | | | | |

### Logistic regression of factors associated with anaemia in paediatric Males living with HIV

In the univariable analysis, increasing age was associated with lower odds of anaemia (OR = 0.79; 95% CI: 0.71-0.88; *p* < 0.0001), indicating that younger children were more likely to be anaemic. Children who received any cotrimoxazole (CTX) during treatment had higher odds of anaemia (OR = 2.07; 95% CI: 1.06-4.01; *p* = 0.031), whereas those with unknown CTX status had lower odds of anaemia (OR = 0.31; 95% CI: 0.12–0.80; *p* = 0.016). Additionally, higher creatinine levels were slightly associated with reduced odds of anaemia (OR = 0.97; 95% CI: 0.95-0.99; *p* = 0.031).

After adjusting for other factors, age remained significantly associated with anaemia. Each additional year of age reduced the odds of anaemia (AOR = 0.81; 95% CI: 0.69- 0.96; *p* = 0.015), confirming that younger children are at higher risk. The associations observed with CTX use and creatinine levels in the univariable analysis were no longer statistically significant after adjustment (AOR = 3.04; *p* = 0.060 and AOR = 0.97; *p* = 0.075, respectively). See table 3

**Table 3.**
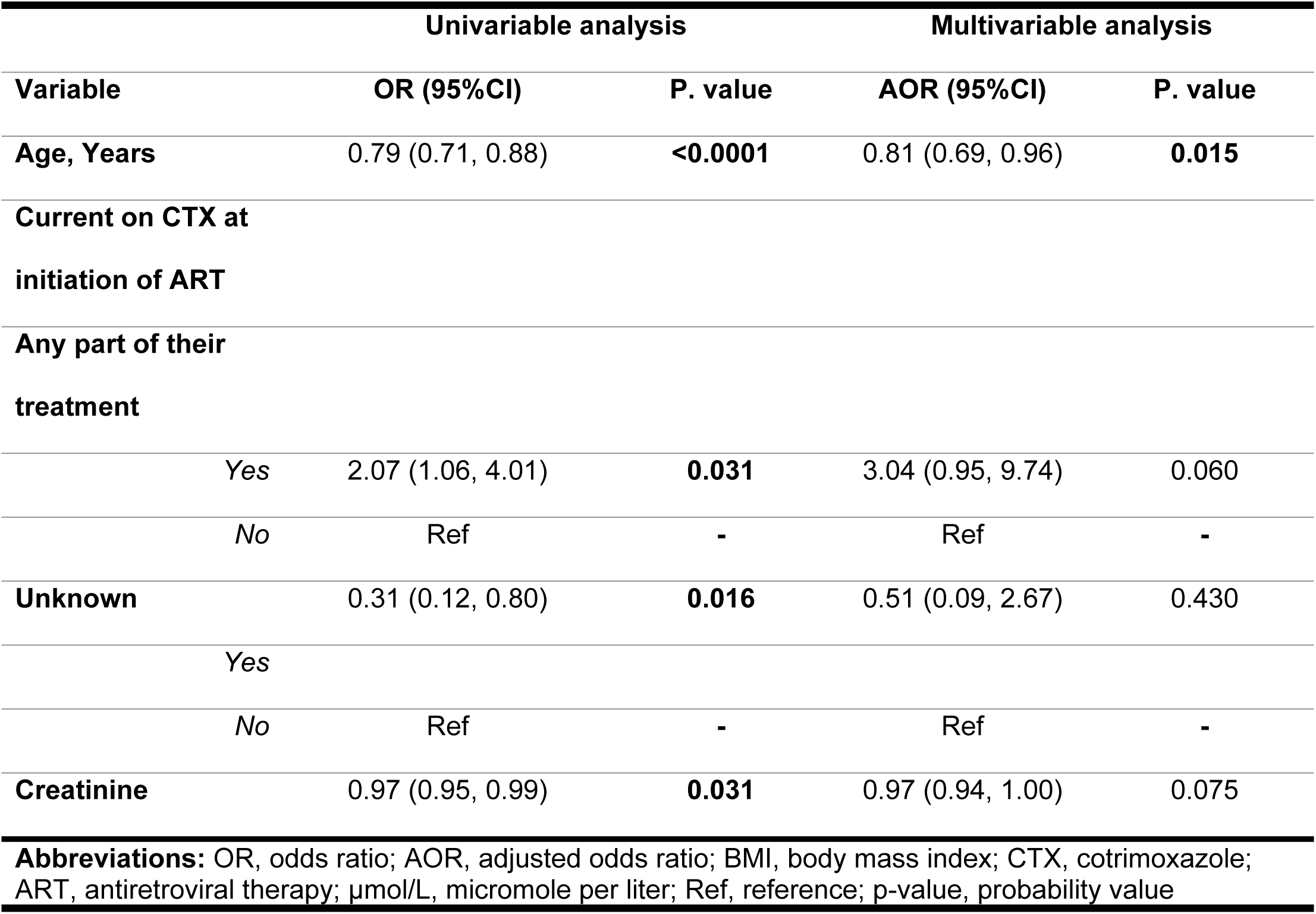
Logistic regression of factors associated with anaemia in CLHIV.

### Logistic regression of factors associated with anaemia in paediatric females living with HIV

In the univariable analysis, increasing age was associated with lower odds of anaemia (OR = 0.82; 95% CI: 0.74– 0.90; p < 0.0001), indicating that younger children were more likely to be anaemic. Children whose primary caregiver was their mother had higher odds of anaemia (OR = 2.46; 95% CI: 1.22–4.96; p = 0.012), whereas those cared for by an aunt had markedly lower odds of anaemia (OR = 0.08; 95% CI: 0.01– 0.66; p = 0.019). Some categories of maternal medication ingestion also showed significant associations, with children in category 3 (OR = 0.20; 95% CI: 0.06–0.67; p = 0.009) and category 4 (OR = 0.31; 95% CI: 0.12– 0.80; p = 0.015) having lower odds of anaemia compared to the reference.

After adjusting for other factors in the multivariable analysis, age remained significantly associated with anaemia (AOR = 0.83; 95% CI: 0.75 – 0.92; p = 0.001), confirming that younger children are at higher risk. Among caregiver variables, being cared for by an aunt remained significantly associated with reduced odds of anaemia (AOR = 0.10; 95% CI: 0.01–0.90; p = 0.040). The associations observed with maternal medication ingestion were no longer statistically significant after adjustment. See table 4

**Table 4.** Logistic regression of factors associated with anaemia in CLHIV.

| Variable | Univariable analysis |  | Multivariable analysis |  |
| --- | --- | --- | --- | --- |
|  | OR (95%CI) | P. value | AOR (95%CI) | P. value |
| **Age, Years** | 0.82 (0.74, 0.90) | **<0.0001** | 0.83 (0.75, 0.92) | **0.001** |
| **Care giver, Mother** |  |  |  |  |
| Yes | 2.46 (1.22, 4.96) | **0.012** | 1.30 (0.58, 2.90) | 0.520 |
| No | Ref | - | Ref | - |
| **Care giver, Aunt** |  |  |  |  |
| Yes | 0.08 (0.01, 0.66) | **0.019** | 0.10 (0.01, 0.90) | **0.040** |
| No | Ref | - | Ref | - |
| **(Mother ingested**<br>**meds)** |  |  |  |  |
| 1 | Ref | - | Ref | - |
| 2 | 0.42 (0.14, 1.20) | 0.106 | 0.58 (0.18, 1.87) | 0.369 |
| 3 | 0.20 (0.06, 0.67) | **0.009** | 0.40 (0.10, 1.50) | 0.176 |
| 4 | 0.31 (0.12, 0.80) | **0.015** | 0.45 (0.16, 1.30) | 0.143 |

**Abbreviations:** OR, odds ratio; AOR, adjusted odds ratio; BMI, body mass index; PMTCT, prevention of mother-to-child transmission; Ref, reference; p-value, probability value.

## Discussion

This study investigated the prevalence and sex-specific determinants of anaemia among children living with HIV (CLHIV) in Zambia’s Southern Province. The findings from our study reveal a substantial burden of anaemia, with a prevalence of 47.0% among children living with HIV in this cohort. Comparable findings have been observed in Malawi (43%), Uganda (46%), and Kenya (49%), underscoring the persistent burden of anaemia in paediatric HIV care [18,19]. Notably, anaemia disproportionately affected younger children, as evidenced by a median age of 8 years in the anaemic group compared to 11 years in the non-anaemic group, pinpointing a younger age as a key determinant of susceptibility. This finding aligns with regional trends in Sub- Saharan Africa (SSA), where anaemia remains a persistent comorbidity despite advances in antiretroviral therapy (ART). For instance, a study by Teklemariam et al., conducted in Ethiopia reported concordant findings, demonstrating that male children had a significantly [20]. Similarly, Beletew et al. from the same setting observed a comparable pattern, attributing the increased susceptibility of younger children to anaemia to the combined effects of elevated physiological iron requirements during rapid growth, inadequate dietary intake, and the high frequency of recurrent infections that typify early childhood [21]. Chronic immune activation and systemic inflammation suppress bone marrow function, while direct viral invasion and infiltration of the marrow further impair red blood cell production [22,23]. Additionally, opportunistic infections such as tuberculosis along with nutritional deficiencies and medication-related toxicity, compound this suppression, leading to persistent and often severe anaemia in advanced stages of HIV disease [24,25].

Although the overall prevalence did not differ significantly by sex, male children exhibited a higher frequency of anaemia (52.6%) compared to females (41.9%)[26]. This pattern aligns with findings from a Tanzanian cohort, where boys demonstrated greater vulnerability to hematologic suppression, potentially due to sex-linked differences in erythropoiesis and iron metabolism [19]. This finding contrasts with general population trends but aligns with evidence from paediatric HIV cohorts, suggesting boys may experience greater nutritional vulnerability and differential immune modulation in the context of chronic infection [21,27]. These findings collectively highlight the multifactorial pathogenesis of anaemia in CLHIV, arising from a complex interaction of age, biological sex, and the underlying chronic infection. These interrelated factors likely disrupt haematopoiesis, nutritional balance, and immune regulatory pathways, could ultimately influencing individual susceptibility to anaemia and its clinical severity [28,29].

Our study also revealed that Increasing age was a consistent and independent protective factor against anaemia in this cohort of CLHIV. The analysis revealed a significant inverse relationship, where each additional year of age was associated with lower odds of anaemia. Notably, anaemia disproportionately affected younger children, as evidenced by a median age of 8 years in the anaemic group compared to 11 years in the non-anaemic group, pinpointing a younger age as a key determinant of susceptibility. This finding aligns with established evidence from across sub-Saharan Africa. Particularly in a study conducted in Mali, their study also revealed that younger children living with HIV were at a significantly higher risk of developing anaemia compared to their older counterparts [30]. Similarly, a study conducted in Botswana reported concordant findings, showing that younger females were particularly vulnerable to anaemia, thereby reinforcing the association between younger age and increased susceptibility within paediatric HIV populations [31].

Maternal caregiving and maternal PMTCT exposure were notable predictors, particularly among female children. This may reflect differential caregiving dynamics or earlier ART exposure in female infants through PMTCT programs [32,33]. Maternal caregivers may also influence feeding patterns and adherence, which could indirectly affect hematologic outcomes [34]. Among male children, cotrimoxazole (CTX) use during treatment appeared as a more prominent determinant of anaemia. This association may indicate a higher burden of concurrent infections among boys, leading to more frequent or prolonged CTX exposure. Furthermore, the drug’s potential to induce oxidative stress and haemolysis in erythrocytes could further contribute to the observed susceptibility to anaemia in this group [34,35].

### Strengths

This study provides several important contributions to the understanding of anaemia in children living with HIV (CLHIV). A key strength is its explicit focus on sex-based differences, moving beyond the traditional approach of treating children as a homogeneous group. By employing sex-stratified analyses, we uncovered distinct risk profiles that would have been obscured in a pooled analysis. For instance, the strong protective association of aunt caregiving was specific to female children, while the trend towards harm with cotrimoxazole (CTX) was specific to males. This nuanced evidence is crucial for developing targeted, sex-sensitive clinical interventions and public health strategies. Another significant strength is the use of programmatic data from a large, real-world cohort across multiple health facilities in Southern Zambia. This enhances the generalizability of our findings to similar sub-Saharan African settings and reflects the outcomes achievable within routine healthcare systems, thereby increasing the practical relevance of our conclusions for policy makers and clinicians.

### Limitations

Despite the strengths there are some limitations which where encountered. The cross-sectional nature of our analysis precludes the determination of causality. We can identify associations but cannot definitively establish whether the identified factors caused anaemia or are merely correlated with it. Another major limitation is reflected in the wide confidence intervals observed for some key estimates, particularly in the multivariable models. For example, the adjusted odds ratio for CTX use in males was 3.04 with a 95% CI of 0.95 to 9.74, and the AOR for aunt caregiving in females was 0.10 with a CI of 0.01 to 0.90. These wide intervals indicate a substantial degree of statistical uncertainty. They are likely a consequence of the relatively small sample size when stratified by sex, which reduces the precision of our estimates and statistical power to detect significant associations. Furthermore, we lacked data on several important potential confounders. Information on dietary iron intake, micronutrient levels (e.g., ferritin, vitamin B12, folate), the specific presence of malaria or helminth infections, and detailed socioeconomic status was not available. Their absence means that residual confounding could influence the observed associations. For instance, the protective effect of aunt caregiving could be linked to unmeasured socioeconomic or nutritional advantages. Finally, the use of data from a single province in Zambia, while a strength for internal consistency, may limit the direct extrapolation of our findings to other geographic and cultural contexts where caregiving dynamics and health system factors may differ. Despite these limitations, the identification of striking sex-specific determinants provides a critical foundation for re-evaluating paediatric HIV care paradigms and underscores the necessity of incorporating a gender lens into both clinical practice and future research.

## Conclusion

This study confirms the high prevalence of anaemia (affecting nearly half the cohort) among children living with HIV in Zambia and, more importantly, revealed distinct sex-specific determinants. A higher burden in males was linked to factors like cotrimoxazole use, suggesting unique vulnerabilities. In females, a powerful protective effect was found with aunt caregiving, highlighting the role of psychosocial factors. These divergent pathways necessitate a shift from a one-size-fits-all approach to gender-sensitive clinical practice. This includes tailored interventions like intensified monitoring for boys on prophylaxis and strengthening support systems for girls. While wider confidence intervals indicate a need for validation in larger studies, the clear signal remains that biological sex is a critical modifier of anaemia risk. Future research should be prospective and include data on micronutrients and socio-economics. Ultimately, responding to these disparities is an ethical and practical imperative for achieving equitable health outcomes for all children living with HIV

## Data Availability

The raw data underlying the results presented in the study have been uploaded as supporting information.

## Author’s contributions

MC and SKM conceived the Study. SKM, BMH and MC oversaw Data acquisition. SKM, MC, BMH and LS supervised data acquisition. SKM, DC and MC conducted the formal analysis. MC, LS, DC, BMH and SKM wrote the original draft. All authors contributed to the article edits and approved the final manuscript.

## Supplementary files

S1. Strobe checklist

S2. Data

S3. Figure 1. Sample size flow chart

